# Nudge strategies for behavior-based prevention and control of neglected tropical diseases: a scoping review and ethical assessment

**DOI:** 10.1101/2020.12.22.20248692

**Authors:** Fiona Vande Velde, Hans J. Overgaard, Sheri Bastien

**Affiliations:** Department of Public Health Science, Faculty of Landscape and Society, Norwegian University of Life Sciences, Ås, Norway; Faculty of Science and Technology, Norwegian University of Life Sciences, Ås, Norway; Department of Microbiology,Faculty of Medicine, Khon Kaen University, Khon Kaen, Thailand; Department of Community Health Sciences, Cumming School of Medicine, University of Calgary, Calgary, Canada; The Centre for Evidence-Based Public Health: A JBI Affiliated Group, Department of Public Health Science, NMBU

## Abstract

**Background:** Nudging, a strategy that uses subtle stimuli to direct people’s behavior, has recently been included as effective and low-cost behavior change strategy in low- and middle- income countries (LMIC), targeting behavior-based prevention and control of neglected tropical diseases (NTDs). Therefore, the present scoping review aims to provide a timely overview of how nudge interventions have been applied within health promotion research, with a specific focus on the prevention and control of NTDs. In addition, the review proposes a framework for the ethical reflection of nudges for behavior-based prevention and control of NTDs, or more broadly global health promotion.

**Methods:** A comprehensive search was performed in the following databases: MEDLINE, PsycINFO, and Embase (Ovid), Web of Science Core Collection, CINAHL, ERIC and Econ.Lit (EBSCO), as well as registered trials and reviews in CENTRAL and PROSPERO to identify ongoing or unpublished studies. Additionally, studies were included through a handpicked search on websites of governmental nudge units and global health or development organizations. A PRISMA flow diagram was used to elaborate on the number of articles retrieved, retained, excluded and reasons for every action.

**Results:** This scoping review of studies implementing nudge strategies for behavior-based prevention and control of NTDs identified 33 studies and a total of 67 nudge-type interventions. Most nudges targeted handwashing behavior and were focused on general health practices rather than targeting a disease in specific. The most common nudge techniques were those targeting decision assistance, such as facilitating commitment and reminder actions. The ethical assessment presented favorable results, certainly regarding the health benefits of the included nudges and the trust relationship for the implementers.

**Conclusion:** Two key recommendations that should inform future research when implementing nudge strategies in global health promotion in general. Firstly, aim for the application of robust study designs including rigorous process and impact evaluation which allow for a better understanding of ‘what works’ and ‘how it works’. Secondly, consider the ethical implications of implementing nudge strategies, specifically in LMIC.

## Introduction

The United Nations formally recognized and included neglected tropical diseases (NTDs) in the Sustainable Development Goals (SDGs) with the following targets: “By 2030, end the epidemics of AIDS, tuberculosis, malaria and neglected tropical diseases and combat hepatitis, water-borne diseases and other communicable diseases” (SDG 3.3) [1]. Currently, preventive chemotherapy (i.e. low-cost mass drug administration without individual diagnosis) is at the core of most NTD programs [2]. However, mass drug administration relies heavily on stable health care infrastructure, which is not always available [3], and additionally, high therapeutic selection pressure could result in drug resistance and eventually lead to unsustainable control [4]. Therefore, a holistic approach to combating NTDs in the long term is widely recommended. This implies shifting away from the focus on medical approaches to an integrated and multisectoral approach [5]. The BEST (Behavior, Environment, Social inclusion, and Treatment) framework was developed by the NTD Non-Governmental Organization Network (NNN) to allow for a comprehensive approach towards NTDs [6]. Nevertheless, comprehensive- and integrated-approaches, although more effective in achieving sustained control, tend to be more resource demanding [7]. However, there is an underinvestment in research concerning NTDs, reflecting a pervasive inequality in global health financing [3]. Therefore, there is an urgent need for effective, low-cost interventions targeting NTDs, stretching beyond the current therapeutic focus.

Recent studies in handwashing behavior for NTD prevention and control present evidence for “nudging” or similar alternative approaches as effective and low-cost behavior change strategies [8]. The purpose of nudging is to subtly direct people towards positive behavioral choices. Nudging does not preclude people’s ability to choose, but instead subtly steers people to make certain favorable decisions for themselves or others. Favorable decisions or favorable behavior relating to health should have a positive health outcome for the people or patients involved [9]. Nudging is grounded in behavioral economics, which is a discipline combining both economics and psychology, and aims to provide an alternative perspective to the assumption that behavior is governed by rational decision-making, as exemplified in traditional economics [10]. The theory acknowledges the limitations inherent to human decision-making, and identifies the cause as our “bounded rationality”; i.e. the limitation of human rationality by several factors such as cognitive and emotional biases, peer and time pressure, among other factors [11]. Thaler and Sunstein [12] address these limitations by presenting the nudge as a behavioral strategy, making the insights from behavioral economics more applicable and accessible. According to the authors, who do not offer a definitive definition but merely suggest an interpretation of the term, a nudge is “…any aspect of the choice architecture (i.e., the design of different ways in which choices can be presented) that alters people’s behavior in a predictable way without forbidding any options or significantly changing their economic incentives. To count as a mere nudge, the intervention must be easy and cheap to avoid” [12] p.6. The definition of a nudge has since been updated to provide further clarity and to reconcile with its theoretical underpinnings in the behavioral sciences [13]. Moreover, Thaler and Sunstein’s definition of a nudge is difficult to operationalize, as it states only that nudges lead to predictable change in human behavior and are different from significant economic incentives or regulation.

In this scoping review, we use the updated definition suggested by Hansen [14] (p.174): “A nudge is a function of an attempt at influencing people’s judgment choice or behavior in a predictable way, that is (1) made possible because of cognitive boundaries, biases, routines, and habits in individual and social decision-making posing barriers for people to perform rationally in their own self-declared interest, and which (2) works by making use of those boundaries, biases, routines, and habits as integral parts of such attempts. Thus a nudge amongst other things works independently of: (i) forbidding or adding any rationally relevant choice options, (ii) changing incentives, whether regarded in terms of time, trouble, social sanctions, economic and so forth, or (iii) the provision of factual information and rational argumentation.”

Nudges have been found to be effective in promoting health without limiting people’s freedom [15], nevertheless, the approach has been widely criticized due to its paternalistic nature. A nudge assumes individuals are not rational actors, capable of making more favorable decisions, which defers responsibility to experts and those in a position of power [16]. Moreover, it questions an individual’s autonomy of choice [17]. However, Thaler and Sunstein [18] use the term “libertarian paternalism,” which underlines the freedom of choice, whilst attributing some responsibility to the nudger (i.e., the person or group instigating the nudge). Considering nudge interventions for targeting NTDs, which mainly affect low- and middle-income countries (LMIC) [19], and largely concentrated among the poorest populations [20], it is imperative to be aware of ethical considerations. Nevertheless, even if power dynamics are at play, some researchers encourage the implementation of nudges, as long as these are suitably transparent and democratically controlled [21].

The present review aims to provide a timely overview of how nudge interventions have been applied within health promotion research, with a specific focus on the prevention and control of NTDs, and what the results have shown thus far. Additionally, in order to inform future efforts directed at implementing nudge interventions, we present a set of ethical criteria that can guide the development of health promotion strategies. In line with these aims and given the relatively recent inclusion of nudges targeting infectious diseases, we opted for a scoping review methodology. This scoping approach allows us to summarize and map the available evidence in nudges for NTD-related research, and additionally, to evaluate relevant ethical considerations that should be taken into account when implementing nudges. The objectives of this scoping review are to: (1) map existing studies which apply a nudge strategy within an intervention for the prevention and control of NTDs in LMIC, (2) identify knowledge gaps to inform future research, (3) propose a framework for the ethical consideration of nudges for NTD prevention and control.

## Methods

In order to ensure a transparent and systematic approach we utilized the JBI Reviewer’s Manual methodology for scoping reviews [22], and the Preferred Reporting Items for Systematic reviews and Meta-Analyses extension for Scoping Reviews (PRISMA-ScR) checklist [23] for reporting. We did not aim to systematically assess the quality of the available studies, required for systematic literature reviews, nor were we concerned with the effectiveness of the nudge strategies. We have focused on providing a broad overview of the field of nudging for the prevention and control of NTDs, implemented in LMIC. Due to the interdisciplinary nature of the topic and the relatively recent emergence of nudging as a strategy, a broader approach was preferred over a systematic quality assessment. In that sense, scoping reviews are particularly useful since they bring together literature from diverse disciplines, and with different approaches to health, intervention, and measurement outcomes. To date there have not been any systematic reviews of any kind in the peer reviewed literature which have focused on nudge strategies for infectious disease control, and more specifically prevention and control of NTDs [24].

### Search strategy

Recommendations for scoping reviews suggest that the search strategy be as inclusive and comprehensive as possible. Since the term “nudging” was only recently established and operationalized, studies with a similar focus could potentially be excluded from the review due to different terms used to describe the nudging technique. The challenge of missing eligible studies because of inconsistent labeling of the term “nudge” has been reported previously [25]. Therefore, we considered including relevant and alternative terms of nudging, as well as sub-categories of the technique. To identify and include these terms, we used a similar approach as Möllenkamp et al. [26]. In addition, we aimed to capture all nudge strategies related to prevention and control of NTDs, and not merely targeting these diseases specifically. Therefore, nudge strategies aimed at changing individual or general health-specific behaviors (e.g. handwashing behavior or non-NTD vector control), although not specifically focused on preventing or controlling a particular type or group of NTDs, were also included in the search strategy. The search algorithm is presented in the study protocol [24], and the final search strategy in MEDLINE presented in Appendix I.

A comprehensive search in the following databases was performed: MEDLINE, PsycINFO, Embase (Ovid), Web of Science Core Collection, CINAHL, ERIC and Econ.Lit (EBSCO), as well as registered trials and reviews in CENTRAL and PROSPERO to identify in-progress or unpublished studies related to the scope of the review. Moreover, references of included studies and literature reviews were screened (based on “Criteria and screening procedure” below), as well as a citation tracking in Web of Science and Google Scholar. In addition, we searched for grey literature on websites of governmental nudge units, such as the Behavioral Insights Team (UK), the Social and Behavioral Science Team (US), the Behavioral Economics Team (AU), the Ministry of Manpower (SG), and iNudgeyou (DK), but also the World Health Organization (WHO) and WHO trial repository, the World Bank (WB) and WB open knowledge repository, United States Agency for International Development (USAID), the Organization for Economic Co-operation and Development (OECD) and OECD library, the United Nations, Abdul Latif Jameel Poverty Action Lab (Jpal), BVA global and BVA Nudge Unit, Busara Center for Behavioral Economics, The International Initiative for Impact Evaluation (3ie), globalhandwashing.org, and behaviouralscientist.org.

### Selection criteria and screening procedure

Articles were screened based on pre-defined selection criteria and procedures [24]. However, the criteria were adapted due to the nature of the literature and deviated to some extent from the protocol, which is described throughout the methods section. The final eligibility criteria are presented in Table 1. We included all full-text papers and reports, peer reviewed articles and gray literature in English. Initially, no restrictions were placed on the type of evidence or study design. However, these inclusion criteria were adapted fairly rapidly through the process, since an outcome measurement of the intervention was required for the data extraction. Review articles were excluded, but screened for identification of missed studies that were subsequently added to the database of included studies. Conference abstracts, as well as editorial letters and comments, were excluded from further analysis. Additionally, to be eligible for inclusion, the nudge strategy had to be described in full, excluding all studies that had an incomplete description of the intervention. All interventions including a nudge strategy were reviewed, regardless of whether they included the explicit label of a “nudge” or not. Initially, to count as a nudge we considered the description by Thaler and Sunstein [12], however, this quickly resulted in an excessive amount of studies. Thaler and Sunstein’s definition of a nudge is difficult to operationalize, as it states only that nudges lead to predictable change in human behavior and are different from significant economic incentives or regulation. Therefore, we consulted an expert in nudging, and adopted the more delimited definition of nudging proposed by Hansen [14]. Thus, articles describing a type of intervention strategy that did not qualify as a nudge based on the definition by Hansen [14] were excluded. Finally, no adaptations were made to the population and context criteria, which included all populations exposed to a nudge strategy targeting behavioral practices for NTD prevention and control in LMIC. All contexts were considered eligible, e.g. public spaces, health care facilities, school settings, and indoor/outdoor community facilities.

**Table 1.** Final eligibility criteria

| Inclusion criteria | Exclusion criteria |
| --- | --- |
| Full-text papers and reports, peer reviewed articles and gray literature | Literature reviews, conference abstracts, editorial letters and comments, theoretical/background papers |
| Written in English language | Written in other languages than English |
| Context of the study is low- and middle- income countries | Context of the study is high-income countries |
| Targeted all behavioral practices leading to NTD prevention and control | Targeted other types of behavioral practices leading to prevention and control of other diseases |
| Behaviorally informed intervention strategies attributed to a nudge strategy, regardless of this explicit label | Interventions not attributed to a nudge strategy, according to the definition by Hansen (2016) |
| The nudge intervention described in full | An incomplete description of the nudge strategy |
| Study design makes it possible to isolate the effect of the nudge on health/implementation outcomes | Study design does not allow for the measurement of the effect of the nudge |

All literature was downloaded by one review author (FVV) to EndNote X9 (Clarivate Analytics, PA, USA), and duplicates removed. The titles and abstracts were screened by one reviewer only (FVV), due to the large number of results. Eligible studies were selected through a questionnaire that specified the inclusion criteria. Two reviewers (SB and FVV) subsequently reviewed the full-text studies independently from each other. Any disagreements concerning eligibility were resolved through discussion. In addition, two independent experts in nudging were contacted to resolve disagreement to eventually reach a consensus.

### Data extraction

All papers selected for inclusion were subjected to a data extraction procedure designed by one review author (FVV) and agreed upon by the other review authors. The extraction form included the following predefined categories: authors; journal/source; year of publication; type of publication; geographical area of the intervention; setting of the intervention; domain of preventive practices (e.g. hygiene, vector control); targeted behavior description (e.g. handwashing); targeted NTD, if specified (e.g. dengue); targeted population; protection of self or others (i.e. whether the behavior is focused on protecting oneself such as through handwashing, or other behaviors such as vector control in the community); nudge strategy; underlying informative theory; and intervention results. Moreover, the nudge strategies were categorized using a choice architecture taxonomy developed by Münscher et al. [27] (Table 2).

**Table 2.** Choice architecture categories and techniques by Münscher et al., 2016

| Category | Description | Technique | Examples |
| --- | --- | --- | --- |
| A. Decision information | Presenting decision-relevant information without changing existing options | A1 Translate information | Message reframing, simplifying |
|  |  | A2 Make information visible | Feedback on own behavior, accessibility of external information |
|  |  | A3 Provide social reference point | Refer to a descriptive norm, or an opinion leader |
| B. Decision structure | Designing or changing options and associated consequences | B1 Change choice defaults | No-action default, prompted choice |
|  |  | B2 Change option related effort | Increase/decrease of physical/financial effort |
|  |  | B3 Change range or composition of options | Change category/group of options |
|  |  | B4 Change option consequences | Change benefit/cost/social consequences of the decision |
| C. Decision assistance | Supporting existing intentions to change | C1 Provide reminders | Un-/materialized reminders |
|  |  | C2 Facilitate commitment | Support self- or public commitment |

In addition, given that nudges raise ethical concerns, the interventions were evaluated based on an adapted ethical framework created by Engelen [28]. To bridge the gap between the abstract theoretical debates among academics and the actual behavioral interventions being implemented in practice, Engelen developed a set of criteria to facilitate an ethical evaluation of different types of nudges [28]. The corresponding categories, criteria and coding labels were adapted to fit the scope of the review, as well as the scope of the included studies, and are presented in Table 3. The original framework presented nine ethical criteria that are categorized into three larger groups: criteria for ends, i.e. evaluation of people’s goals and values; criteria for means, i.e. evaluation of people’s decision-making process; criteria for agents i.e. evaluation of the role of the nudgers. In the current scoping review, the categories are reflected accordingly: criteria for targeted behaviors, which are considered the ‘ends’ or the result of the implemented strategy; criteria for interventions, which relate to the nudge strategy and process in itself; criteria for researchers, the agents responsible for implementing the study. The ethical criteria were subsequently adapted to ensure relevance and facilitate the assessment on the included studies. We aimed at adhering to the original criteria, but this was not always possible due to the limited information reported in the manuscripts. Three criteria changed categories: low processing motivation; democratic legitimation; easy resistibility. For example, ‘low processing motivation’ became a criterion for the targeted behaviors, since we were not able to deduce the way the intervention (e.g., the means) was processed by the participants, based on the included information. However, the studies included information on the repetitiveness and importance of the targeted behavior (e.g., habitual handwashing, taking care of infants), hence the necessity of performing the behavior through a high-or low-cognitive route. Extracting this information allowed us to develop coding labels, and subsequently evaluate the criterion. A similar process supported the development of all other criteria. The criterion ‘rational capacities’ was excluded from the assessment, since we were unable to identify relevant coding labels that fitted the studies. Finally, we developed three evaluation codes to indicate the degree to which studies met each criteria: High (H); Moderate (M); Low (L).

**Table 3.** Criteria for ethical assessment of nudge strategies targeting health promotion behaviors related to NTDs in LMIC. Adapted from Engelen (2019).

| Category | Criteria | Description | Label | Code<sup>1</sup> |
| --- | --- | --- | --- | --- |
| 1. Criteria for targeted behaviors | Reflective preferences | Nudges should be based on people's own reflective preferences | Targeted behavior is underpinned by population preferences | H |
|  |  |  | Formative research conducted, but not used to target the behavior | M |
|  |  |  | No mention of formative research or behavioral preferences | L |
|  | Health benefits | Nudges should generate greater health benefits | Health benefits are immediate by implementing the behavior | H |
|  |  |  | Health benefits depend on the involvement of the community | M |
|  |  |  | Health benefits depend on many other environmental factors | L |
|  | Low processing motivation | Nudges should engage in low processing motivation | Behavior is repetitive, low processing is needed | H |
|  |  |  | Behavior is implemented during certain moments, or for a certain group | M |
|  |  |  | Behavior is performed only once, therefore high stakes | L |
| 2. Criteria for interventions | Democratic legitimization | Nudges should gain broad public support | Intervention is developed through participatory approaches or iterations | H |
|  |  |  | Intervention is with insights from formative research or by a local agency | M |
|  |  |  | Intervention is not developed with community reflection or feedback | L |
|  | Easy resistibility | Nudges should allow people with opposite preference to go against them | Awareness of the nudge and other options are within reach | H |
|  |  |  | Awareness of the nudge, but other options are fairly unreachable | M |
|  |  |  | No awareness of the nudge and no other options available | L |
|  | Long-run autonomy | Nudges should generate greater autonomy in the long run | Both short- and long- run autonomy are preserved | H |
|  |  |  | No short-, but long- run autonomy is preserved | M |
|  |  |  | Both short- and long- run autonomy are not preserved | L |
|  | Available alternatives | Nudges should present more effective than information or persuasion | Nudge showed a positive outcome, more effective than alternatives | H |
|  |  |  | No adequate experimental design, but study showed a positive outcome | M |
|  |  |  | Nudge showed a negative outcome, less effective than alternatives | L |
| 3.Criteria for researchers | Trust relationship | Nudges should be implemented by agents in a trust relationship with the nudges | One of the authors affiliated with a local institution | H |
|  |  |  | No authors affiliated with a local institution, but includes local collaborators | M |
|  |  |  | No mention of local collaboration throughout the study | L |

<sup>1</sup>H = high ethical standard; M = moderate ethical standard; L = low ethical standard

The ethical assessment was developed concurrently with the data extraction form and both were pilot tested to minimize misinterpretation and to ensure all relevant data were included in the analysis. Given the diversity of studies, the piloting was conducted among a sample of five studies comprising different domains of preventive practices and with diverse study designs. The piloting was performed by one review author through several iterations (FVV) and discussed at length with a second reviewer (SB). This resulted in the inclusion of several categories in the data extraction form: authors’ affiliations; behavioral intervention package complexity (i.e. largest number of intervention components implemented to a group participants; simple: 1-2, moderate: 3-4, and complex: >4); theoretical underpinning or design process of the whole intervention package (e.g. Evo-Eco theory, RANAS); nudge materials (e.g., posters, painted cues, messages); research design; secondary outcomes related to the nudge; intervention outcomes attributed to the nudge; experimental design. Ultimately, the data extraction procedure was conducted by one review author (FVV), whilst a second reviewer (SB) validated 25% of the extracted information. Disagreements were resolved through discussion until consensus was reached.

## Results

### Study selection

The search strategy resulted in a total of 2497 records retrieved from the specified databases, with 1792 records remaining after removal of duplicates (Figure 1). The selection process was iterative as the criteria were developed based on the discussions amongst the review authors and experts’ inputs, and became more exclusive and bounded to improve conceptual clarity. In the first full-text examination round, one review author (FVV) sorted the 169 selected records into ‘include’, ‘exclude’ or ‘uncertain’ selection. The ‘uncertain’ selection was discussed with a second review author (SB), and eventually synthesized into three main topics of uncertainty: emotional framing, low-cost innovations, and mental visualizations. We reached out to two experts to assist with resolving the uncertainty associated with these approaches. Additionally, one expert provided additional premises and guidelines: a nudge is an aspect of an intentional intervention, not the intervention itself; the intervention can therefore include both rational aspects as well as nudges at the same time; a nudge does not affect action by provision of rational reasoning; actions can also include mental events (e.g. belief-formation, direction of attention); simply creating an opportunity does not count as a nudge; fear of social sanctions does not count as a nudge; and lastly, a nudge implemented to alter the mental or emotional state should be supported by evidence and not merely assumed. Subsequently, the eligibility criteria were modified, uncertain records were examined for inclusion, and previously excluded records based on ‘ineligible intervention’ were re-assessed. After an extensive search for potentially missed studies and grey literature, concluded on September 6^th^, 2020, two independent reviewers (SB and FVV) initiated a second round of full-text examination on a total of 68 records. Eventually, and with far less discussion, this resulted in 33 identified studies.

**Figure 1.**
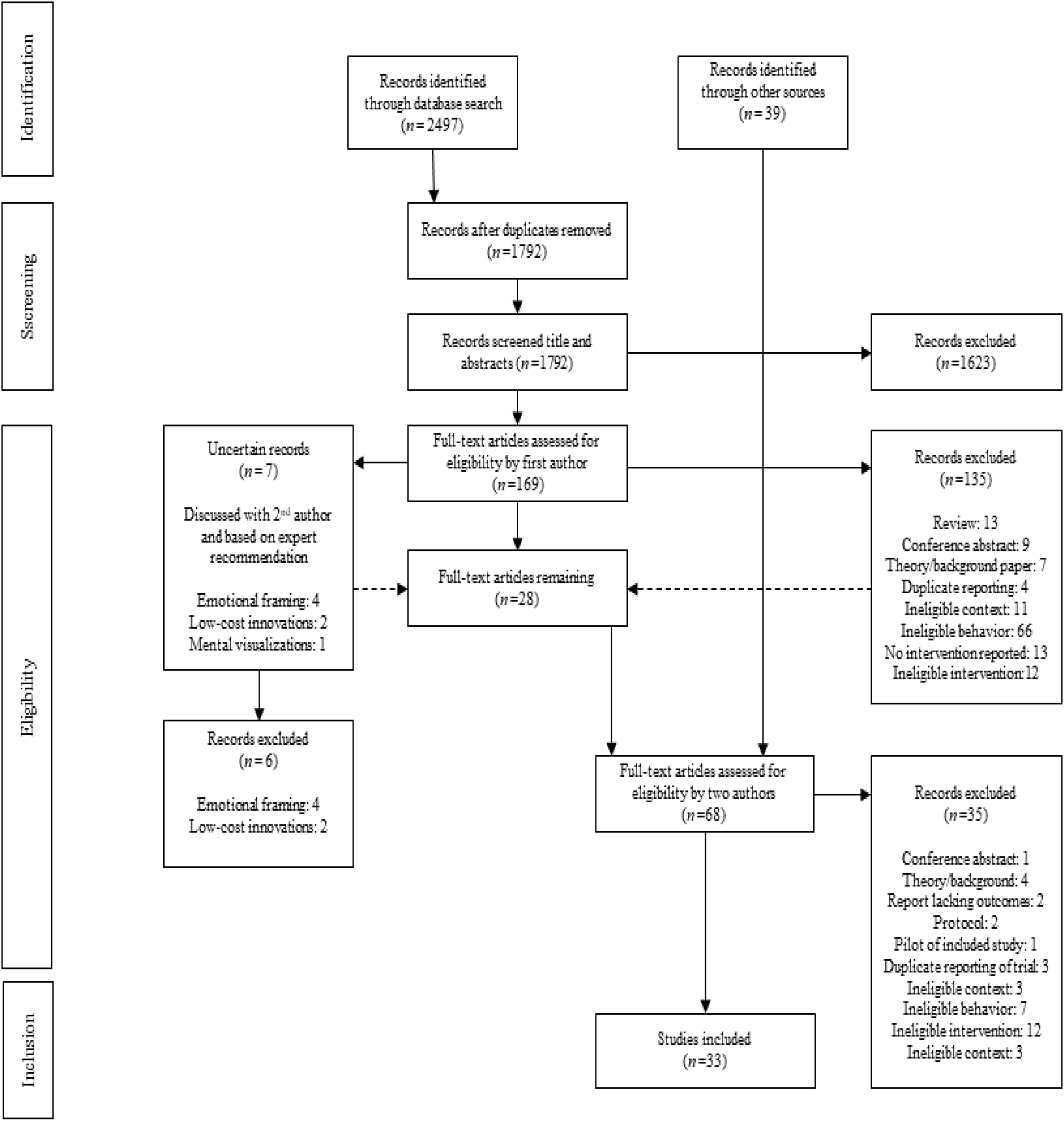
Flow diagram based on the Preferred Reporting Items for Systematic reviews and Meta-Analyses extension for Scoping Reviews (PRISMA-ScR) checklist (Tricco et al., 2018). Note that the selection process was iterative and resulted in reconsideration and subsequent inclusion of studies for eligibility assessment (dashed lines).

### Notable eligibility decisions

Despite the application of stringent eligibility criteria in an objective and systematic fashion, there is unavoidably an element of subjectivity involved, and certainly when considering the complexity of the nudge definition. In an attempt to make this subjectivity transparent, this sub-section reports on the eligibility decisions made. These decisions led to the selection of a full study, or merely the selection of an intervention if the study implemented multiple nudge strategies within the intervention.

According to the eligibility criteria, the nudge intervention had to be described in full, excluding all studies that had an incomplete description of the intervention. This criterion is closely associated with the expert’s premise that a nudge is an aspect of an intentional intervention, and not the intervention itself. Due to this statement, we excluded a number of studies that implemented a ‘reminder’ as an aspect of the intervention, without explicitly describing the process the reminder was targeting. Merely describing the instrument (e.g., poster, song) was not sufficient to be included in the analysis. Some studies implemented posters or murals as reminders to instigate a certain preventive behavior, but failed to describe the precise location for this reminder to be activated (e.g. posters near the sink to activate handwashing action). Interventions that mentioned ‘posters were placed in a strategic location’ [29] or even developed and implemented through participatory processes [30] were further excluded from analysis if they lacked sufficient detail as to the context and location for the nudge. Interventions implementing songs or radio advertisements as reminders were also excluded from the analysis, since the timing of these interventions was not controlled and therefore it was not possible to be considered as a reminder [31].

Targeting of social norms was applied in many interventions and studies, however, such an approach must be used in a certain manner in order to be labeled a nudge. The norms should be descriptive (e.g., what people do), rather than injunctive (e.g., what people should do), since the latter induces fear of social sanctions. Subsequently, avoiding social sanctions is regarded as a rational decision and is, therefore, not a nudge. Any form of peer pressure or promotion of injunctive norms were excluded from further analysis. For example, an intervention on infant feeding in Vietnam based on gossip induced fear of social sanctions among young mothers if they did not implement adequate infant feeding practices [32]. Another aspect of exclusion was based on inadequate research design. If the study design did not allow for deducing the effect of the social norms, due to missing secondary outcomes or insufficient evidence of the intervention being linked to social constructs, the intervention was deemed ineligible for further analysis [33].

Similar to the latter argument presented for social norms, we excluded interventions that aimed at prompting action by altering decision information through certain cognitive or emotional processes without explicitly measuring this hypothesized cognitive or emotional model. Several interventions based on emotional triggers such as disgust and nurture were not included due to a lack of empirical information about the intervention processing [34, 35]. We were not able to infer whether the action was instigated by merely a matter of information provision or whether participants were exposed to a nudge. So called ‘triggering’ motivating behavior change by activating a collective sense of disgust [36], mainly used in community-led total sanitation efforts, were equally subjected to this exclusion argument.

The provision of tippy taps, i.e. locally made low-cost devices for washing hands with running water [37] or other ‘enabling technologies’, were not considered eligible interventions. The majority of these interventions merely provided an opportunity, which eventually created the desired action (i.e. handwashing). In addition, these interventions can arguably be categorized as some form of frugal and innovative sanitary engineering, which also falls beyond the scope of this review.

### Study characteristics

As detailed previously, 33 papers were retained following full-text analysis [38-70]. This section will present the eligible records and the extracted data relating to the study characteristics. The complete list of citations and extracted data presented below are available in Appendix II. The full extracted data is available on request from the first author.

#### Date of publication

All included records were published since 2010 with the number of studies increasing exponentially, with a substantial rise from 2017 onwards. During that first modest increase (2010 – 2017), 12 studies were published [39, 41, 46, 49, 56, 59-61, 63-65, 67]. The exponential increase is most noticeable in 2018 resulting in 7 eligible studies [42, 43, 48, 50, 55, 57, 58], while in 2019, there were 9 included studies [38, 44, 45, 52, 54, 62, 66, 69, 70], and finally up until 6^th^ September 2020 we identified 5 additional studies [40, 47, 51, 53, 68].

#### Study locations

Half of the included studies were conducted in Asia (16 records), with India making up the largest cohort of studies [39, 44, 47, 48, 57, 63, 68], followed by Bangladesh [38, 41, 50, 59], the Philippines [51, 54] Nepal [49], and Iraq [69]. One study was conducted in both Bangladesh and Kenya [60]. The other half of the included studies were conducted in Africa (15 records): four studies in Kenya [52, 55, 66, 70], followed by two in Zimbabwe [53, 56], Nigeria [40], South-Africa [42], Malawi [45], Ethiopia [46], Chad [58], Burundi [61], Zambia [62], Mali [64], Egypt [65], and Uganda [67]. Finally, only one study was identified which took place on the South-American continent: Peru [43].

#### Setting and participants

Half of the studies were conducted in a rural setting (16 records [39-41, 44-49, 52, 53, 55, 58, 61, 64, 68]). Eight studies took place in an urban/peri-urban setting, of which 6 studies specified a low-income/slum context [38, 42, 59, 60, 63, 67], the remaining 2 did not specify the context of the setting [43, 56]. A total of 8 studies specifically targeted schools [50, 51, 54, 57, 62, 65, 66, 70], and one study focused on a humanitarian emergency setting [69].

The participants included only mothers or caregivers of children under the age of 5 (5 records [38, 46, 47, 49, 58]), both caregivers and children (2 records [57, 63]), and only including children (8 records [42, 50, 51, 54, 62, 66, 69, 70]). Several studies targeted an entire household (8 records [39-41, 43, 47, 48, 60, 68]) or the entire community (7 records [44, 53, 55, 56, 59, 61, 64]), whilst a fewer records specified targeting the school community [65], young women [52], or users of a certain facility with specified traits such as shared compound toilets [67].

#### Targeted behavior

The included studies did not specify a disease which the intervention was focused on specifically, apart from two studies targeting Chagas disease [43] and trachoma [66]. The studies targeted specific behaviors rather than diseases and merely described possible health effects of these behaviors, mostly a decrease of infectious pathogens causing a host of illnesses (e.g., diarrhea, pneumonia). Half of the studies targeted handwashing with soap (HWWS) or simply handwashing (17 records [38-42, 46, 50, 51, 53, 54, 57, 59, 62, 63, 65, 69, 70]). Seven studies targeted defecation behaviors such as latrine usage, building or cleaning, and safe disposal of child feces [44, 47, 48, 61, 64, 67, 68], 4 studies focused on water disinfection through chlorination or solar disinfection [52, 56, 58, 60], 2 studies targeted a mix of food hygiene behaviors [45, 49], one study each targeted drug intake for deworming [55], face washing [66], and engaging the population to participate in an indoor residual spraying (IRS) campaign [43].

#### Intervention strategies

A range of intervention strategies were employed across the included studies, with some studies either detailing multiple, combined behavior change strategies (including nudges), others incorporating few strategies, whilst some merely focused on applying one strategy. Half of the eligible studies consisted of a complex intervention (17 records [39, 40, 44, 45, 47, 49, 53, 54, 56-59, 63-66, 68]). Subsequently, a total of 7 ‘moderate intervention’ studies were recorded [41, 43, 46, 48, 51, 60, 70]. Finally, 9 records consisted of a simple intervention [38, 42, 50, 52, 55, 61, 62, 67, 69].

### Nudge characteristics

We identified a total of 67 nudges in 33 studies, this section presents the extracted nudges based on the targeted behavior, the different categories of choice architecture, and the ethical standards.

#### Nudges and targeted behavior

Thirty of the nudges were implemented for HWWS in a total of 17 studies [38-42, 46, 50, 51, 53, 54, 57, 59, 62, 63, 65, 69, 70] (Figure 2). This group of nudges were most diverse and included a range of behavior change strategies, such as the implementation of disruptive cues: colorful, painted footpaths [50, 51]; soap including embedded toys [42, 69]; soap on a rope [62]; posters as reminders [51, 59, 65]; stickers as reminders [39, 41, 51], the implementation of social norms: stickers or certificates as signals of pro-social behavior [39, 40, 59]; role-model display [39]; and public pledges [39, 40, 46, 53, 54, 57, 63, 70] by simplifying information [38] and providing planning reminders [53]. Twelve nudges targeted defecation behaviors in 7 studies [44, 47, 48, 61, 64, 67, 68]. Most of these studies deployed commitment as a strategy, e.g. self-commitment [44, 61] or public pledges [47, 64, 67, 68]. Others included social norms such as signaling (banners [44], family picture [47]); role models [48]; public competitions [64], but also by providing sticker reminders [47], and by providing visual feedback in coloring the village open defecation “hot spots” [44]. Disinfecting water was targeted by 11 nudges in 4 studies [52, 56, 58, 60]: signaling (stickers [56] or ribbons [60]), public pledges [58, 60], message framing [58, 60], reminder stickers [56], simplifying the behavior through planning [52], or visualizing the future [52]. Only two studies focused on food hygiene behaviors, but these included a total of 7 nudges: public pledges [45, 49], signaling (stickers [45] and ‘safe food’ zones [49]), competitions, colorful reminders in the kitchen and role-model display [49]. One study targeted vector control and participation in an IRS campaign for Chagas’ disease control [43], including 4 nudges: planning commitment, associated planning-reminders, role-models promoting participation, and contingent group lotteries. Another study included two nudges for deworming behavior with social signaling through bracelets and reminder messages [55]. Finally, one nudge targeted face washing through a public pledge in a classroom setting [66]. Figure 2 presents the percent of the nudges distributed per behavior.

**Figure 2.**
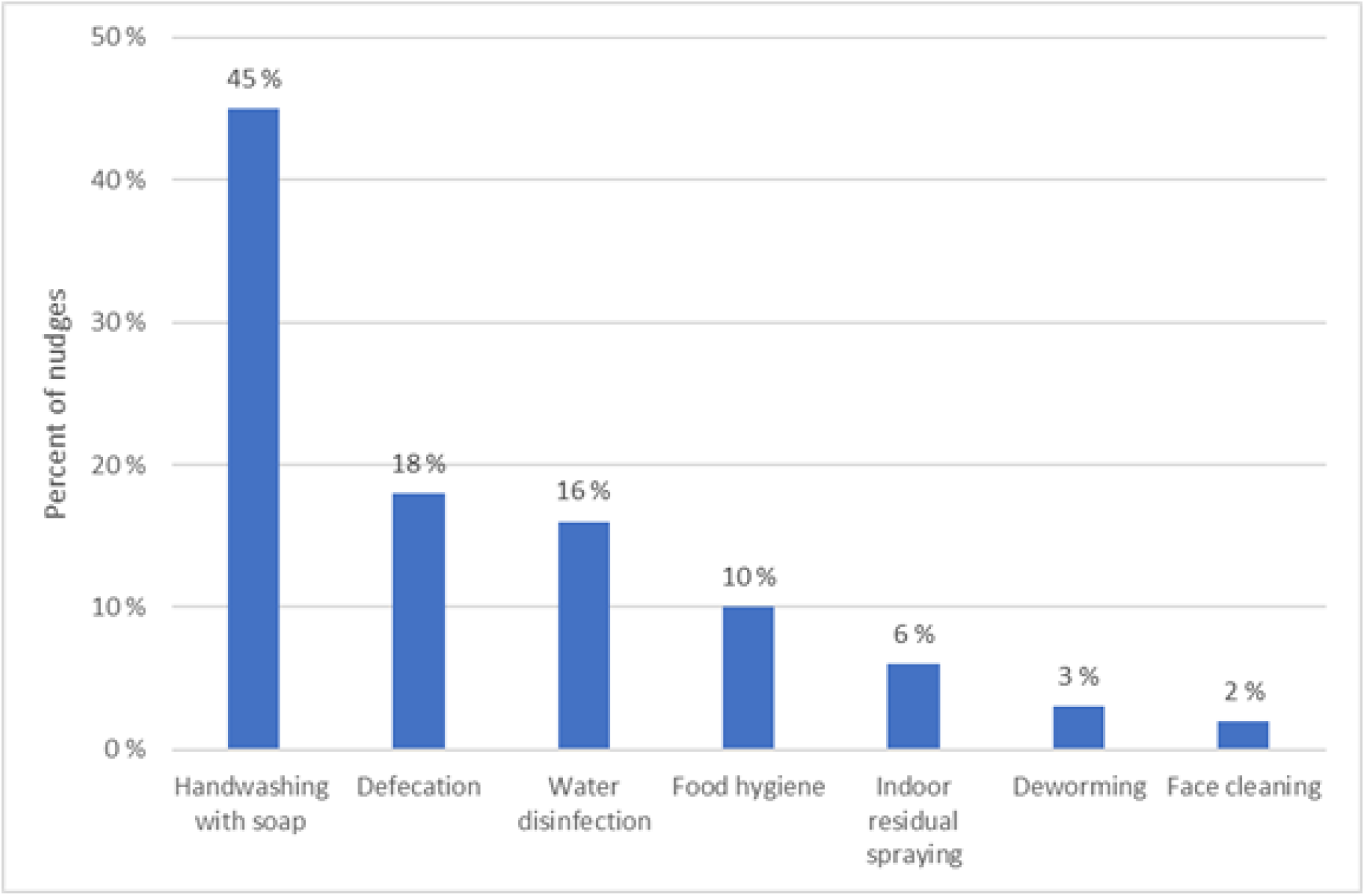
Percent of nudges included within the review (n = 67) that targeted: a. Handwashing with soap (HWWS) (30); b. Defecation behaviors (12); c. Water disinfection (11); d. Food hygiene (7); e. Indoor residual spraying (IRS) (4); f. Deworming (2); g. Face cleaning (1)

#### Choice architecture

The included nudge strategies were categorized into different techniques (Table 2) and depicted in Figure 3. The most common nudge techniques were those targeting decision assistance, such as “facilitating commitment” (24 nudges), representing both self-commitment (including planning commitment) and public commitment (public pledges) [39, 40, 43-47, 49, 53, 54, 57, 58, 60, 61, 63, 64, 66-68, 70] and “providing reminders” (16 nudges), using posters, stickers, and planning reminders [39, 41, 43, 47, 49-51, 53, 55-57, 59, 62, 65]. Techniques targeting decision structure represented by “change option consequences” only, included 16 nudges using signaling, competitions and lotteries (including toy-soap, since the chances of ‘winning’ the toy depends on the amount of soap used) [39, 40, 42-45, 47, 49, 55, 56, 58, 59, 64, 69]. Finally, decision information was the least used category with 5 nudges using “translate information” through simplification and message reframing [38, 52, 58, 60], 3 nudges using “provide social reference point” referring to role-models [39, 43, 48, 49], and 2 nudges implementing “make information visible” through visualization of the future and providing visual feedback [44, 52].

**Figure 3.**
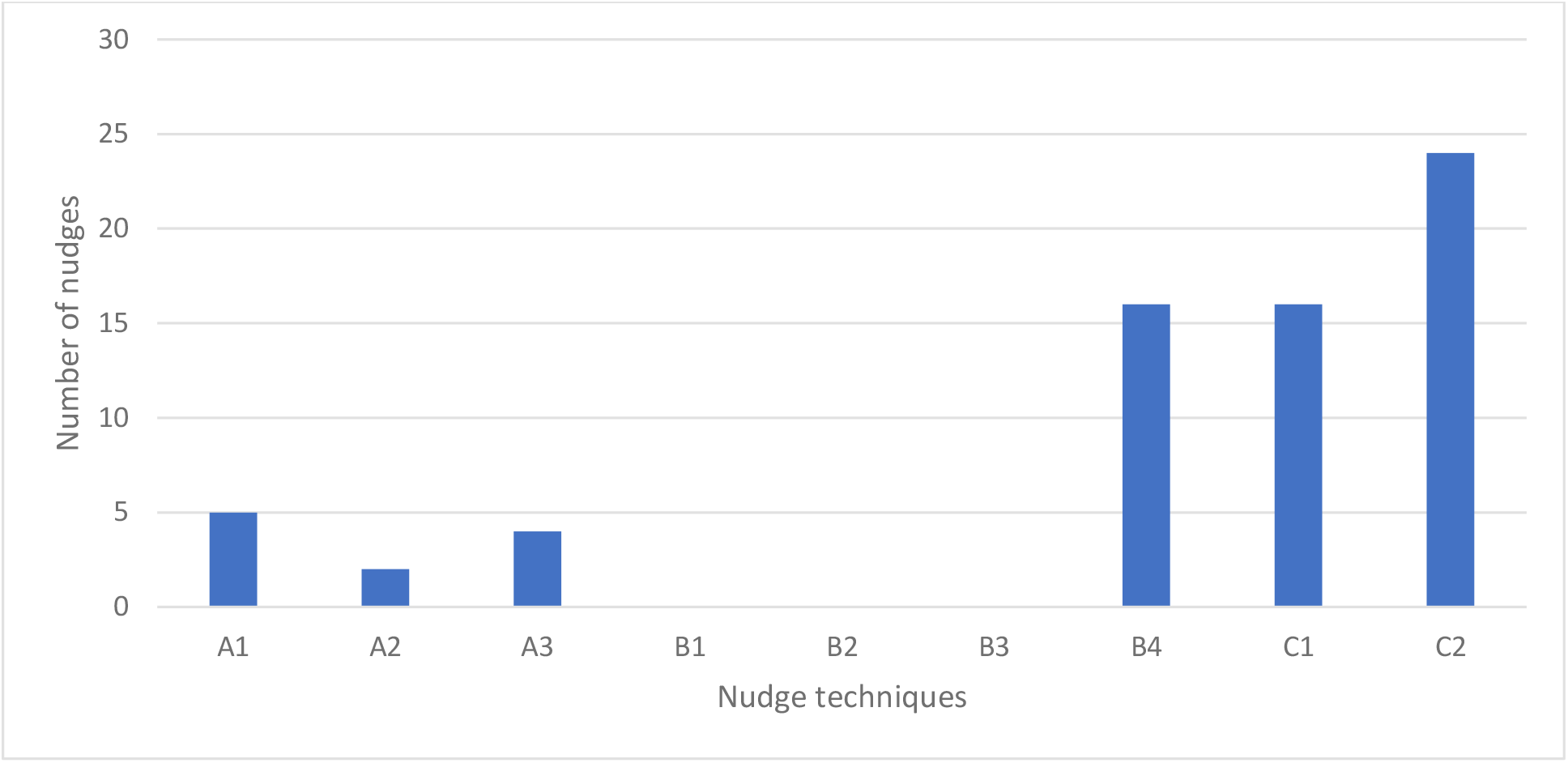
Number of identified nudge strategies in selected studies based on the three choice architecture categories A. Decision information, B. Decision structure, and C. Decision assistance (Münscher et al. 2016). A1 Translate information; A2 Make information visible; A3 Provide social reference point; B1 Change choice defaults; B2 Change option related effort; B3 Change range or composition of options; B4 Change option consequences; C1 Provide reminders; C2 Facilitate commitment

#### Ethical assessment

The results of each of the eight ethical criteria described in Table 3 are presented below one by one, and the findings are visualized in Figure 4, through the corresponding codes (i.e., H = high; M = moderate; L = low – ethical standard). A complete list of citations and scoring can be found in Appendix III.

**Figure 4.**
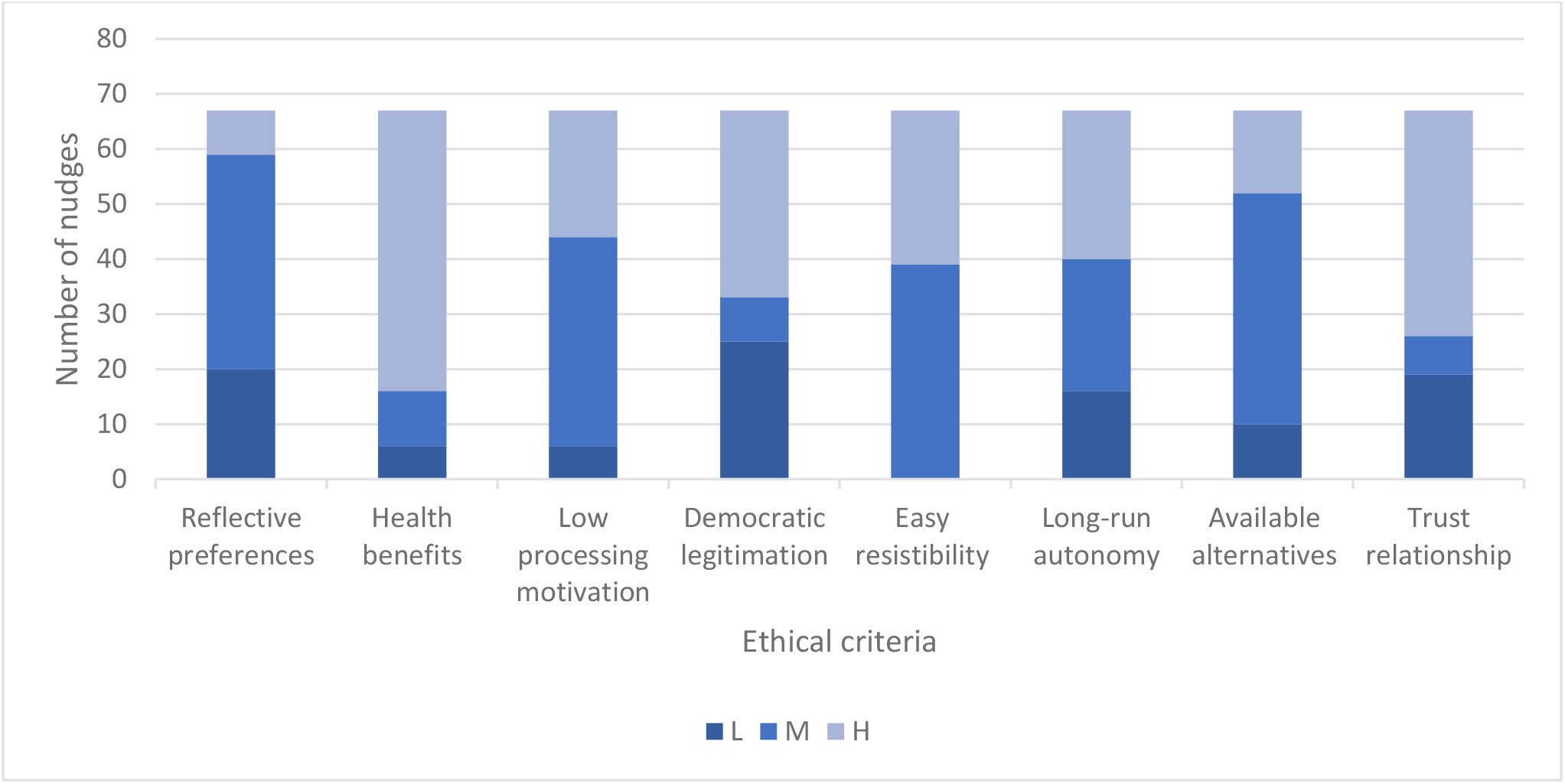
Ethical assessment of 67 nudge strategies targeting health promotion behaviors related to NTDs in LMICs. Nudge strategies assessed according to eight criteria: Reflective preferences (RP); Health benefits (HB); Low processing motivation (LP); Democratic legitimation (DL); Easy resistibility (ER); Long-run autonomy (LA); Available alternatives (AA); Trust relationship (TR) were categorized as being of a high (H), moderate (M), and low (L) ethical standard.

“Reflective preferences” were assessed by extracting the data regarding formative research: H, 8 nudges were developed through behavioral preferences of the targeted population (e.g. identifying irrelevant behavior to target together with the community); M, 39 nudges were developed through pre-defined behavior, but with some formative knowledge of the population (e.g. assessing the barriers or norms to a certain behavior); L, 20 nudges were developed without any research on population’s preferences (e.g. no mention of formative research).

“Health benefits” were assessed based on whether the targeted behavior results in immediate positive health outcomes or to what degree these health outcomes depended on other factors apart from the behavior: H, 51 nudges had immediate health benefits (e.g. HWWS); M, 10 nudges depend on community participation (e.g. defecation behaviors); L, 6 nudges depend on other environmental factors, beyond the targeted behavior (e.g. cleaning latrines).

“Low-processing motivation” is assessed through the repetitiveness of the targeted behavior, as well as the magnitude of the impact: H, 23 nudges targeted low-processing behaviors (e.g. HWWS); M, 38 nudges targeted behaviors during specified moments such as water disinfection, or involving a certain group (e.g. infant feeding); L, 6 nudges were targeting one-time, key decisions (e.g. deworming). The second category “criteria for means” is concerned with the implemented nudge strategy.

“Democratic legitimation” is assessed through extraction of the data regarding the development of the intervention: H, 34 nudges were developed with the community (e.g. co-creation of intervention materials); M, 8 nudges were based on formative research, but without direct community involvement (e.g. local design agency); L, 25 nudges were designed by the research team without local involvement or knowledge (e.g. materials were pre-designed).

“Easy resistibility” is based on the awareness of the nudge and whether other options (or opting out) are visible and within reach of the nudgee: H, 28 nudges were highly visible and easy to avoid if the participant preferred to not engage (e.g. reminders); M, 39 nudges were also visible, but more difficult to oppose (e.g. all public behaviors); L, no nudges were invisible or hiding other options (e.g. default).

“Long-run autonomy” is based on the assessment of both short-term decisions and modifications during the intervention, and longer-term changes after the study has come to a conclusion: H, 27 nudges secure both short- and long-run autonomy (e.g. reminders); M, 24 nudges affected short-term decisions, but long-run autonomy is preserved (e.g. public commitment); L, 16 nudges make significant changes to the environment and result in both short- and long-run loss of autonomy (e.g. signaling).

“Available alternatives” are assessed through the reported health outcomes of the studies: H, 15 nudges were more effective than the presented alternative options (e.g. adequate experimental design with a positive effect attributed to the nudge); M, 42 nudges were part of a larger intervention set that reported a positive health outcome (e.g. intervention set compared to other or control); L, 10 nudges reported negative effects compared to the alternative (e.g. adequate experimental design with a negative effect attributed to the nudge).

Finally, “trust relationship” consisted of one item assessing the trust relationship between the nudgers and the nudgees, in this case the relationship between the research team and targeted population: H, 41 nudges were implemented by a local collaborator as part of the research team (e.g. minimum of one affiliated author); M, 7 nudges were implemented by a local collaborator that was not part of the research team (e.g. no locally affiliated author); L, 19 nudges were implemented by a foreign research team without mentioning local collaboration (e.g. the study did not mention local involvement).

## Discussion

In this scoping review, we identified a total of 33 studies that included a nudge strategy for behavior-based prevention and control of NTDs in LMIC. Only two studies targeted NTDs in particular, trachoma and Chagas disease, the other 31 included studies were focused on more general health protective behaviors such as handwashing behaviors. Accordingly, handwashing with soap accounted for almost half of the included nudge strategies, followed by defecation behaviors (e.g., latrine usage, cleaning), and water disinfection behaviors. Targeting health behaviors rather than diseases, hence implementing a horizontal-rather than a vertical approach, has increasingly become common practice in global health and health promotion, and specifically concerning WASH-related efforts [71]. This holistic view is also reflected in the complexity of the studies and the inclusion of multi-layered, complex interventions. The majority of the studies implemented interventions that include several different behavior change strategies, which made it particularly difficult to identify nudge strategies in reported studies. Consequently, nudges were difficult to isolate, and eventually to attribute their effectiveness within a complex intervention. Although this was not the purpose of our scoping review, it identifies an important knowledge gap that needs addressing in terms of advancing the field.

Based on Münscher et al. [27] the current study reports category C nudges, ‘decision assistance’, to be most popular in targeting behaviors for NTD prevention and control, followed by category B, ‘nudges targeting decision structure’, and finally category A nudges, ‘altering decision information’. Category C nudges are most transparent and ethical since they merely provide assistance to help nudgees to follow through with their intentions. Commitment nudges (C2) were also commonly applied, and these can be distinguished between self- and public-commitment. However, we did not differentiate these since most self-commitment nudges to some extent involved public commitments, due to at least one witness as part of the research team being present. Reminders (C1) were frequently reported, however, due to the lack of detailed description concerning the context in which they were implemented, we were unable to consider them as nudges.

Category B interventions are considered least transparent or ethical due to hiding or changing other, less favorable options. Noteworthy is that we only found type B4 nudges, change option consequences (social or beneficial/costs), which is to some extent in line with other reviews focused on nudging and health-related behaviors [26, 72]. Distinct from these other reviews, our included studies targeted social consequences/encouragement, rather than connecting these decisions to micro-benefits or costs. One explanation could be that in LMICs, and in particular rural settings, the importance of the community takes greater precedence than the individual. Thereby making this type of nudge more appropriate and strategic to target social consequences/encouragement, such as ‘signaling’ pro-social behaviors.

Finally, nudges based on category A were reported least frequently. Similar to reminders, many of the studies including message framing (e.g., emotions), presentation of social norms, visualizing or simplifying information were excluded from further analysis, due to a lack of detailed description and validation of the process. Triggering negative emotions such as disgust or shame, or positive emotions such as nurture and fun, are frequently used in global health promotion and specifically in community-led total sanitation approaches [36]. However, there is a significant lack of empirical verification of the causality of these emotional processes. We therefore question the use and effect of many of these interventions, as have others [73], which presents another important research gap.

### Identifying areas for future research opportunities

Two major areas for future research were identified through this scoping review, which present an opportunity to learn from and include other research methodologies and fields: (1) ‘what works’; and (2) ‘how it works’. Firstly, the majority of the included studies consisted of a complex set of interventions, which is common practice in health promotion and is largely encouraged [74]. However, many of these studies were not concerned with understanding what specific component of the intervention was effective, a knowledge gap that is presented in other literature reviews [75]. The research designs were primarily basic experimental approaches or longitudinal studies, exposing one group of the population to the intervention versus a control group that did not receive any form of intervention. We suggest that future studies employ more robust research designs, which allow for isolating and investigating the effectiveness of different strategies (such as nudges) which may be nested within a more complex intervention. A stepped-wedge experimental approach, exposing each group gradually to a certain intervention, and measuring changes in behavior over a certain time-period [76] could be an appropriate study design to allow for this. Secondly, a large amount of studies was considered ineligible due to a lack of causality between the proposed behavioral technique (e.g., social norms, emotions, framing) and the behavior. Hence, lacking an understanding on how the nudge strategies were eventually processed and adopted by the participants. We suggest including robust empirical verification of the proposed causality and effects of these behavioral techniques, such as pre-testing of the proposed materials and including secondary outcome measurements in the larger trial. By addressing these two main knowledge gaps, future research will be able to establish the evidence base concerning how nudges work and the effectiveness of different types of nudges in different contexts and among different target groups.

### Proposed ethical framework

The evaluation of nudge strategy ethics was based on the criteria developed by Engelen [28], which were adapted to fit our purpose. We did not aim to develop a novel method for assessing ethical standards more broadly within nudging for global health and health promotion, but merely included this to advance the discussion and debate concerning the ethics of nudges. Therefore, we did not label nudges as ‘ethical’ or ‘unethical’. We simply aimed to provide a general assessment of nudges applied in the reviewed studies. Two criteria stand out, including most H-labeled nudges in their assessment: ‘health benefits’ and ‘trust relationship’. Health benefits can be defined as measured by both the magnitude of the impact and the numbers of people positively affected by performing the behavior instigated by the nudge [28]. Since the scoping review focused on NTDs or related behaviors that prevent infectious disease transmission, hence affecting many people, it is only logical that this criterion would gain much support. However, most studies targeted behaviors rather than specific diseases, which made it difficult to quantify the health benefits. Therefore, we differentiated between these benefits based on their immediate effect and whether these depended on other factors besides the behavior of the nudgee. Nevertheless, the targeted behaviors and associated nudges remained highly beneficial for health, and therefore more legitimate to implement according to Engelen [28]. Trust relationship is the only criterion evaluating the ‘nudgers’, in our case the research team instigating the nudge. For this purpose, we extracted the data based on the inclusion of local collaborators and affiliation of the authors. Most studies included at least one local author, however, to our surprise, some studies did not include or acknowledge collaboration with local institutions or local authorship. Described further in the limitations, we have based this assessment on what is presented in the manuscript and did not make any efforts to contact the authors to clarify inclusion of local partners. Nevertheless, the involvement of local collaborators was assumed rather high, which is considered more ethical for implementing nudges.

‘Reflective preferences’ and ‘Democratic legitimation’, were two criteria which received a lower ethical assessment in the included studies. Both criteria measure the involvement by the local population in developing the study, however, focused on two different outcomes of this participation. Reflective preferences assess the involvement of the population in targeting the behavior and associated needs. Many of the included studies did not include, or only to some extent, formative research to identify and understand the context and culture of the population. Most studies decided in advance which behaviors they targeted from an epidemiological perspective. However, in order to develop more democratically legitimate nudges, formative research should also focus on the preferred behaviors from the perspective of the target population. Democratic legitimation, a criterion targeting the intervention itself, evaluates the cultural reflection for developing the nudge interventions. The criterion indicates rather low ethical standards, which means that the nudge strategies were primarily decided upon by the research team without the involvement of local stakeholders. Developing intervention strategies using a participatory approach, is more likely to result in greater support and endorsement by the population, which may subsequently result in more ethical, legitimate and effective nudges [77].

Overall, the majority of nudges in the assessed studies had moderately to high ethical standards. This positive finding highlights awareness among researchers of the ethical considerations associated with nudge strategies for behavior-based prevention and control of NTDs. We propose the criteria included in the ethical assessment as a relevant guideline for future development and research of nudge strategies in global health promotion in general. Following these proposed guidelines may foster more explicit consideration of the principles suggested by Schmidt [21], which encourages the implementation of nudges as long as these are suitably transparent and democratically controlled. Furthermore, future research could include other types of strategies, similar to nudging, such as boosting [78]. Proponents of both nudging and boosting agree that human decision-making is often deficient, and that these deficits are caused by our bounded rationality. However, boosts and nudges differ mainly in ‘how’ they aim at improving decision-making. While nudges coopt people’s cognitive biases to affect behavioral changes, boosts have been positioned as being more likely to empower and foster individual autonomy and competence to make their own choices, by training people in employing existing decision heuristics or employing new ones [79]. These alternatives could be used in combination with more traditional forms of nudging for global health promotion, identified by this scoping review, and foster population’s health, capacities and resilience in the long run.

## Limitations of the review

### Search strategy

The scoping review was designed to be as inclusive as possible, which resulted in an extensive search and several iterations of study selection. However, the nature of the literature did not allow for a simple search strategy such as others have done [72, 80], where the boundary was set to studies that exclusively use the term “nudge” or “choice architecture”. Interventions that include nudges have only recently been identified as promising strategies for NTD prevention and control, or more broadly infectious disease and global health promotion. By restricting our search to only those manuscripts mentioning these terms, our scoping review would lose much of its value, since few studies would meet this inclusion criteria. Moreover, there has been some tradition in global health promotion to include interventions that fall under the category nudge, although not explicitly labelling it as such (e.g., commitment, message framing). However, many of these studies use different terminologies, such as public commitment or pledge, which made it more complex for the research team to identify the interventions consistently. Nevertheless, other reviews have been reported with a broad scope on nudge-type interventions, however, these have a more targeted behavioral topic, such as the self-management of chronic diseases [26]. We acknowledge that having both a broad scope on the interventions as well as the targeted behavior was highly ambitious, therefore, recognize the possibility of unidentified studies. To overcome this limitation, we published the manuscript as a pre-print, and shared it with the scientific community to allow for comments or identify possible missed studies (medRxiv).

In addition, due to the unforeseen complexity of the literature, we decided to simply focus on English written studies. This has probably led to an underrepresentation of the South-Americas, since research studies and reports are frequently published in Spanish. However, most studies included an English abstract, but many of the interventions which included nudge strategies were only identified through full-text selection rather than title and abstract screening.

### Screening and selection

As discussed in the notable eligibility decisions, there is an unavoidable element of subjectivity involved, and certainly when considering a concept with ‘fuzzy’ boundaries such as nudging. Nevertheless, we believe we have taken steps to be as objective, inclusive and transparent in our reporting as possible. We believe that the scale and nature of our screening and selection approach was appropriate given our emphasis on mapping and collating the existing evidence rather than conducting a full systematic assessment. Moreover, we have recorded and included all steps, as well the modifications to the protocol.

### Methods used for the ethical assessment

The ethical assessment was a first attempt to establish an ethical framework for implementing nudge strategies in behavior-based prevention and control of NTDs. This attempt fit our intended purpose well, but it was not our aim to develop a methodology for assessing the ethics of nudge strategies across all fields. Moreover, some limitations should be taken into account when interpreting the results of this study. Firstly, the ethical assessment was based on what was described in the manuscript of the included studies. Therefore, if the authors did not mention work beyond the scope of their study, or refer to other publications, this was considered in the coding process as being missing. For example, if formative research was not mentioned, or referred to by the authors, this was considered lacking. Similarly, if local collaborators were not mentioned or acknowledged in the text, these were considered not being part of the study. We did not intend to follow up on these issues with the authors, since this was beyond the scope of our assessment. Our aim was to map what is described in the literature, not to conduct research on what could and should have been described. We do acknowledge there might have been some studies incorrectly coded. However, our aim was to present a general overview of the current ethical framework, and guidelines for meeting these ethical standards in future research.

## Conclusions

The main outcomes of this review including 67 nudges for NTD prevention and control can be summarized in two key recommendations that should inform future research when implementing nudge strategies in global health promotion in general. Firstly, aim for the application of robust study designs including rigorous process and impact evaluation which allow for a better understanding of ‘what works’ and ‘how it works’. This will lead to more targeted, and thereby more efficient and effective nudge strategies. Secondly, acknowledge the need for rigorous consideration of the ethical implications of nudge strategies, according to the aforementioned guidelines. Aim for transparency, democracy and autonomy when implementing nudges, specifically in resource constrained settings such as LMIC.

## Supporting information

Appendix III

## Data Availability

The extracted data presented in the manuscript is included as supplementary material in a separate appendix II, in addition, the full extracted data can be obtained through the first author

## Acknowledgements

We would like to thank research librarian Johanne Longva at the Norwegian University of Life Sciences for providing essential support in developing the search strategy and protocol for the scoping review. In addition, we wish to express our gratitude towards the two experts that provided valuable input for resolving eligibility discussions. Furthermore, we thank the members of the Department of Public Health Science (NMBU) for their constructive feedback on the review. Finally, this review was conducted as a part of the MY-SCHOOL project, which received funding from the Research Council of Norway (GLOBVAC Project no. 281588).

## References

1. United Nations: Transforming our world: the 2030 agenda for Sustainable Development. https://sustainabledevelopment.un.org/post2015/transformingourworld, 2015 (accessed 15th October 2020).

2. Webster JP, Molyneux DH, Hotez PJ, Fenwick A: The contribution of mass drug administration to global health: past, present and future. Philos Trans R Soc Lond B Biol Sci 2014, 369(1645):20130434.

3. Molyneux DH, Savioli L, Engels D: Neglected tropical diseases: progress towards addressing the chronic pandemic. The Lancet 2017, 389(10066):312–325.

4. Vercruysse J, Albonico M, Behnke JM, Kotze AC, Prichard RK, McCarthy JS, Montresor A, Levecke B: Is anthelmintic resistance a concern for the control of human soil-transmitted helminths? International Journal for Parasitology: Drugs and Drug Resistance 2011, 1(1):14–27.

5. Freeman MC, Ogden S, Jacobson J, Abbott D, Addiss DG, Amnie AG, Beckwith C, Cairncross S, Callejas R, Colford JM, Jr. et al: Integration of Water, Sanitation, and Hygiene for the Prevention and Control of Neglected Tropical Diseases: A Rationale for Inter-Sectoral Collaboration. PLOS Neglected Tropical Diseases 2013, 7(9):e2439.

6. NNN: The BEST-framework. https://www.ntd-ngonetwork.org/the-best-framework-a-comprehensive-approach-towards-neglected-tropical-diseases, 2019 (accessed 15th October, 2020).

7. Das JK, Salam RA, Arshad A, Maredia H, Bhutta ZA: Community based interventions for the prevention and control of Non-Helmintic NTD. Infect 2014, 3:24–24.

8. Dreibelbis R, Kroeger A, Hossain K, Venkatesh M, Ram PK: Behavior Change without Behavior Change Communication: Nudging Handwashing among Primary School Students in Bangladesh. Int J Environ Res Public Health 2016, 13(1):129.

9. Fridman I, Hart JL, Yadav KN, Higgins ET: Perspectives on using decision-making nudges in physician-patient communications. PLoS ONE 2018, 13(9):e0202874.

10. Camerer C: Behavioral economics: Reunifying psychology and economics. Proceedings of the National Academy of Sciences 1999, 96(19):10575–10577.

11. Kahneman D: A perspective on judgment and choice: mapping bounded rationality. Am Psychol 2003, 58(9):697–720.

12. Thaler RH, Sunstein CR: Nudge: improving decisions about health, wealth, and happiness. New Haven, CT, US: Yale University Press; 2008.

13. Hansen PG: Nudging: To know ‘what works’ you need to know why it works. Journal of Behavioral Economics for Policy 2019, 3 (S):9–11.

14. Hansen PG: The Definition of Nudge and Libertarian Paternalism: Does the Hand Fit the Glove? European Journal of Risk Regulation 2016, 7(1):155–174.

15. Vlaev I, King D, Dolan P, Darzi A: The Theory and Practice of “Nudging”: Changing Health Behaviors. Public Administration Review 2016, 76(4):550–561.

16. Raihani N: Nudge politics: efficacy and ethics. Front Psychol 2013, 4:972.

17. Epstein RA: The Dangerous Allure of Libertarian Paternalism. Review of Behavioral Economics 2018, 5 (3-4):389–416.

18. Thaler RH, Sunstein CR: Libertarian Paternalism. American Economic Review 2003, 93(2):175–179.

19. Stolk WA, Kulik MC, le Rutte EA, Jacobson J, Richardus JH, de Vlas SJ, Houweling TAJ: Between-Country Inequalities in the Neglected Tropical Disease Burden in 1990 and 2010, with Projections for 2020. PLOS Neglected Tropical Diseases 2016, 10(5):e0004560.

20. Houweling TAJ, Karim-Kos HE, Kulik MC, Stolk WA, Haagsma JA, Lenk EJ, Richardus JH, de Vlas SJ: Socioeconomic Inequalities in Neglected Tropical Diseases: A Systematic Review. PLOS Neglected Tropical Diseases 2016, 10(5):e0004546.

21. Schmidt AT: The Power to Nudge. American Political Science Review 2017, 111(2):404–417.

22. Peters MDJ, Marnie C, Tricco AC, Pollock D, Munn Z, Alexander L, McInerney P, Godfrey CM, Khalil H: Updated methodological guidance for the conduct of scoping reviews. JBI Evidence Synthesis 2020, 18(10):2119–2126.

23. Tricco AC, Lillie E, Zarin W, O’Brien KK, Colquhoun H, Levac D, al. e: PRISMA Extension for Scoping Reviews (PRISMA-ScR): Checklist and Explanation. Annals of Internal Medicine 2018, 169(7):467–473.

24. Vande Velde F, Longva J, Overgaard HJ, Bastien S: Identifying nudge strategies for behavior-based prevention and control of neglected tropical diseases: a scoping review protocol. JBI Evidence Synthesis 2020, 18 (12):2704–2713.

25. Dolan P, Hallsworth M, Halpern D, King D, Metcalfe R, Vlaev I: Influencing behaviour: The mindspace way. Journal of Economic Psychology 2012, 33(1):264–277.

26. Möllenkamp M, Zeppernick M, Schreyögg J: The effectiveness of nudges in improving the self-management of patients with chronic diseases: A systematic literature review. Health Policy 2019, 123(12):1199–1209.

27. Münscher R, Vetter M, Scheuerle T: A Review and Taxonomy of Choice Architecture Techniques. Journal of Behavioral Decision Making 2016, 29(5):511–524.

28. Engelen B: Ethical Criteria for Health-Promoting Nudges: A Case-by-Case Analysis. The American Journal of Bioethics 2019, 19(5):48–59.

29. Kitsanapun A, Yamarat K: Evaluating the effectiveness of the “Germ-Free Hands” intervention for improving the hand hygiene practices of public health students. J Multidiscip Healthc 2019, 12:533–541.

30. Graves JM, Daniell WE, Harris JR, Obure AFXO, Quick R: Enhancing a Safe Water Intervention with Student-Created Visual AIDS to Promote Handwashing Behavior in Kenyan Primary Schools. International Quarterly of Community Health Education 2012, 32(4):307–323.

31. Greenland K, Chipungu J, Curtis V, Schmidt WP, Siwale Z, Mudenda M, Chilekwa J, Lewis JJ, Chilengi R: Multiple behaviour change intervention for diarrhoea control in Lusaka, Zambia: a cluster randomised trial. Lancet Glob Health 2016, 4(12):e966–e977.

32. White S, Schmidt W, Sahanggamu D, Fatmaningrum D, van Liere M, Curtis V: Can gossip change nutrition behaviour? Results of a mass media and community-based intervention trial in East Java, Indonesia. Trop Med Int Health 2016, 21(3):348–364.

33. Null C, Stewart CP, Pickering AJ, Dentz HN, Arnold BF, Arnold CD, Benjamin-Chung J, Clasen T, Dewey KG, Fernald LCH et al: Effects of water quality, sanitation, handwashing, and nutritional interventions on diarrhoea and child growth in rural Kenya: a cluster-randomised controlled trial. Lancet Glob Health 2018, 6(3):e316–e329.

34. Mbuya MNN, Tavengwa NV, Stoltzfus RJ, Curtis V, Pelto GH, Ntozini R, Kambarami RA, Fundira D, Malaba TR, Maunze D et al: Design of an Intervention to Minimize Ingestion of Fecal Microbes by Young Children in Rural Zimbabwe. Clinical Infectious Diseases 2015, 61(suppl_7):S703–S709.

35. Humphrey JH, Mbuya MNN, Ntozini R, Moulton LH, Stoltzfus RJ, Tavengwa NV, Mutasa K, Majo F, Mutasa B, Mangwadu G et al: Independent and combined effects of improved water, sanitation, and hygiene, and improved complementary feeding, on child stunting and anaemia in rural Zimbabwe: a cluster-randomised trial. Lancet Glob Health 2019, 7(1):e132–e147.

36. Venkataramanan V, Crocker J, Karon A, Bartram J: Community-Led Total Sanitation: A Mixed-Methods Systematic Review of Evidence and Its Quality. Environ Health Perspect 2018, 126(2):026001.

37. Mbakaya BC, Kalembo FW, Zgambo M: Use, adoption, and effectiveness of tippy-tap handwashing station in promoting hand hygiene practices in resource-limited settings: a systematic review. BMC Public Health 2020, 20(1):1005.

38. Amin N, Sagerman DD, Nizame FA, Das KK, Nuruzzaman M, Yu J, Unicomb L, Luby SP, Ram PK: Effects of complexity of handwashing instructions on handwashing procedure replication in low-income urban slums in Bangladesh: a randomized non-inferiority field trial. Journal of Water, Sanitation and Hygiene for Development 2019, 9(3):416–428.

39. Biran A, Schmidt W-P, Varadharajan KS, Rajaraman D, Kumar R, Greenland K, Gopalan B, Aunger R, Curtis V: Effect of a behaviour-change intervention on handwashing with soap in India (SuperAmma): a cluster-randomised trial. Lancet Glob Health 2014, 2(3):e145–e154.

40. Biran A, White S, Awe B, Greenland K, Akabike K, Chuktu N, Aunger R, Curtis V, Schmidt W, Van der Voorden C: A cluster-randomised trial to evaluate an intervention to promote handwashing in rural Nigeria. International Journal of Environmental Health Research 2020:1–16.

41. Biswas D, Nizame FA, Sanghvi T, Roy S, Luby SP, Unicomb LE: Provision versus promotion to develop a handwashing station: the effect on desired handwashing behavior. BMC Public Health 2017, 17(1):390.

42. Burns J, Maughan-Brown B, Mouzinho A: Washing with hope: evidence of improved handwashing among children in South Africa from a pilot study of a novel soap technology. BMC Public Health 2018, 18(1):709.

43. Buttenheim AM, Paz-Soldan VA, Castillo-Neyra R, Toledo Vizcarra AM, Borrini-Mayori K, McGuire M, Arevalo-Nieto C, Volpp KG, Small DS, Behrman JR et al: Increasing participation in a vector control campaign: a cluster randomised controlled evaluation of behavioural economic interventions in Peru. BMJ glob 2018, 3(5):e000757.

44. Caruso BA, Sclar GD, Routray P, Majorin F, Nagel C, Clasen T: A cluster-randomized multi-level intervention to increase latrine use and safe disposal of child feces in rural Odisha, India: the Sundara Grama research protocol. BMC Public Health 2019, 19(1):322.

45. Chidziwisano K, Slekiene J, Kumwenda S, Mosler H-J, Morse T: Toward Complementary Food Hygiene Practices among Child Caregivers in Rural Malawi. The American Journal of Tropical Medicine and Hygiene 2019, 101(2):294–303.

46. Contzen N, Meili IH, Mosler H-J: Changing handwashing behaviour in southern Ethiopia: A longitudinal study on infrastructural and commitment interventions. Social Science & Medicine 2015, 124:103–114.

47. Friedrich M, Balasundaram T, Muralidharan A, Raman VR, Mosler HJ: Increasing latrine use in rural Karnataka, India using the risks, attitudes, norms, abilities, and self-regulation approach: A cluster-randomized controlled trial. Sci Total Environ 2020, 707:135366.

48. Gauri V, Rahman T, Sen IK: Shifting Social Norms to Reduce Open Defecation in Rural India. In: Policy Research Working Paper; No 8684. vol. Policy Research Working Paper; No. 8684. Washington DC: World Bank; 2018.

49. Gautam OP, Schmidt WP, Cairncross S, Cavill S, Curtis V: Trial of a Novel Intervention to Improve Multiple Food Hygiene Behaviors in Nepal. American Journal of Tropical Medicine and Hygiene 2017, 96(6):1415–1426.

50. Grover E, Hossain MK, Uddin S, Venkatesh M, Ram PK, Dreibelbis R: Comparing the behavioural impact of a nudge-based handwashing intervention to high-intensity hygiene education: a cluster-randomised trial in rural Bangladesh. Tropical Medicine & International Health 2018, 23(1):10–25.

51. Haung C, Le N, Battle M: A nudge toward hand hygiene: simple design features improved handwashing among Filipino students. In. Medium: https://medium.com/idinsight-blog/a-nudge-toward-hand-hygiene-simple-design-features-improved-handwashing-among-filipino-students-ae4fab1c94db: IDInsight; 2020. (accessed June 4th 2020)

52. Haushofer J, John A, Orkin K: Can Simple Psychological Interventions Increase Preventive Health Investment? NBER Working Papers 2019: 25731.

53. Inauen J, Lilje J, Mosler H-J: Refining hand washing interventions by identifying active ingredients: A cluster-randomized controlled trial in rural Zimbabwe. Social Science & Medicine 2020, 245(2020):112712

54. Jetha Q, Villasenor JM, Lehmann LV: Can social motivators improve handwashing behavior among children? Evidence from a cluster randomized trial of a school hygiene intervention in the Philippines, https://www.semanticscholar.org/paper/Title%3A-Can-social-motivators-improve-handwashing-a-Jetha-Villasenor/cdf1f183ae5983573f1b3b10fc71de94999bd249, 2019 (accessed on June 4th 2020)

55. Karing A, Naguib K: The role of social signaling in community deworming: Evidence from a field experiment in Kenya. American Journal of Tropical Medicine and Hygiene 2018, 99 (4 Supplement):173.

56. Kraemer SM, Mosler H-J: Effectiveness and Effects of Promotion Strategies for Behaviour Change: Solar Water Disinfection in Zimbabwe. Applied Psychology 2012, 61(3):392–414.

57. Lewis HE, Greenland K, Curtis V, Schmidt WP: Effect of a School-Based Hygiene Behavior Change Campaign on Handwashing with Soap in Bihar, India: Cluster-Randomized Trial. Am J Trop Med Hyg 2018, 99(4):924–933.

58. Lilje J, Mosler H-J: Effects of a behavior change campaign on household drinking water disinfection in the Lake Chad basin using the RANAS approach. Sci Total Environ 2018, 619-620:1599–1607.

59. Luby SP, Kadir MA, Yushuf Sharker MA, Yeasmin F, Unicomb L, Sirajul Islam M: A community-randomised controlled trial promoting waterless hand sanitizer and handwashing with soap, Dhaka, Bangladesh. Tropical Medicine & International Health 2010, 15(12):1508–1516.

60. Luoto J, Levine D, Albert J, Luby S: Nudging to Use: Achieving Safe Water Behaviors in Kenya and Bangladesh. Journal of Development Economics 2014, 110:13–21.

61. Mosler H-j, Sonego IL: Improved latrine cleanliness through behaviour change and changes in quality of latrine construction: a longitudinal intervention study in rural Burundi. International Journal of Environmental Health Research 2017, 27(5):355–367.

62. Naluonde T, Wakefield C, Markle L, Martin A, Tresphor C, Abdullah R, Larsen DA: A disruptive cue improves handwashing in school children in Zambia. Health Promotion International 2019, 34(6):e119–e128.

63. Nicholson JA, Naeeni M, Hoptroff M, Matheson JR, Roberts AJ, Taylor D, Sidibe M, Weir AJ, Damle SG, Wright RL: An investigation of the effects of a hand washing intervention on health outcomes and school absence using a randomised trial in Indian urban communities. Tropical Medicine & International Health 2014, 19(3):284–292.

64. Pickering AJ, Djebbari H, Lopez C, Coulibaly M, Alzua ML: Effect of a community-led sanitation intervention on child diarrhoea and child growth in rural Mali: a cluster-randomised controlled trial. Lancet Glob Health 2015, 3(11):e701–e711.

65. Talaat M, Afifi S, Dueger E, El-Ashry N, Marfin A, Kandeel A, Mohareb E, El-Sayed N: Effects of hand hygiene campaigns on incidence of laboratory-confirmed influenza and absenteeism in schoolchildren, Cairo, Egypt. Emerg Infect Dis 2011, 17(4):619–625.

66. Tidwell JB, Fergus C, Gopalakrishnan A, Sheth E, Sidibe M, Wohlgemuth L, Jain A, Woods G: Integrating Face Washing into a School-Based, Handwashing Behavior Change Program to Prevent Trachoma in Turkana, Kenya. The American Journal of Tropical Medicine and Hygiene 2019, 101(4):767–773.

67. Tumwebaze IK, Mosler H-J: Effectiveness of group discussions and commitment in improving cleaning behaviour of shared sanitation users in Kampala, Uganda slums. Social Science & Medicine 2015, 147:72–79.

68. Viswanathan S, Saith R, Chakraborty A, Purty N, Malhotra N, Singh P, Mitra P, Padmanabhan V, Datta S, Harris J et al: Improving households’ attitudes and behaviours to increase toilet use (HABIT) in Bihar, India. 3ie Impact Evaluation Report. New Delhi: International Initiative for Impact Evaluation (3ie), 2020, 118.

69. Watson J, Dreibelbis R, Aunger R, Deola C, King K, Long S, Chase RP, Cumming O: Child’s play: Harnessing play and curiosity motives to improve child handwashing in a humanitarian setting. International Journal of Hygiene & Environmental Health 2019, 222(2):177–182.

70. Wichaidit W, Steinacher R, Okal JA, Whinnery J, Null C, Kordas K, Yu J, Pickering AJ, Ram PK: Effect of an equipment-behavior change intervention on handwashing behavior among primary school children in Kenya: the Povu Poa school pilot study. BMC Public Health 2019, 19(1):647.

71. Freeman MC, Stocks ME, Cumming O, Jeandron A, Higgins JPT, Wolf J, Prüss-Ustün A, Bonjour S, Hunter PR, Fewtrell L et al: Systematic review: Hygiene and health: systematic review of handwashing practices worldwide and update of health effects. 2014, 19(8):906–916.

72. Forberger S, Reisch L, Kampfmann T, Zeeb H: Nudging to move: a scoping review of the use of choice architecture interventions to promote physical activity in the general population. International Journal of Behavioral Nutrition and Physical Activity 2019, 16(1):77.

73. Leontsini E, Winch PJ: Increasing handwashing with soap: emotional drivers or social norms? Lancet Glob Health 2014, 2(3):e118–e119.

74. White S, Thorseth AH, Dreibelbis R, Curtis V: The determinants of handwashing behaviour in domestic settings: An integrative systematic review. International Journal of Hygiene and Environmental Health 2020, 227:113512.

75. De Buck E, Van Remoortel H, Hannes K, Govender T, Naidoo S, Avau B, Veegaete AV, Musekiwa A, Lutje V, Cargo M et al: Approaches to promote handwashing and sanitation behaviour change in low- and middle-income countries: a mixed method systematic review. 2017, 13(1):1–447.

76. Haushofer J, Metcalf CJE: Which interventions work best in a pandemic? Science 2020, 368(6495):1063–1065.

77. Hagman W, Andersson D, Västfjäll D, Tinghög G: Public Views on Policies Involving Nudges. Review of Philosophy and Psychology 2015, 6(3):439–453.

78. Hertwig R, Grüne-Yanoff T: Nudging and Boosting: Steering or Empowering Good Decisions. Perspectives on Psychological Science 2017, 12(6):973–986.

79. Grüne-Yanoff T: Boosts vs. Nudges from a Welfarist Perspective. Revue d’économie politique 2018, 128(2):209–224.

80. Hofmann B, Stanak M: Nudging in screening: Literature review and ethical guidance. Patient Educ Couns 2018, 101(9):1561–1569.

