## Appendix I for "Nudge strategies for behavior-based prevention and control of neglected tropical diseases: a scoping review and ethical assessment"

| Medline | Mesh | Free keywords |
| --- | --- | --- |
| String 1 | exp Economics, Behavioral/ | (behavior?r* adj1 economic*).tw,kf. |
|  |  | (nudg* or nudging).tw,kf. |
|  |  | (choice adj1 (architect* or intervention*)).tw,kf. |
|  |  | (behavio?r* adj2 intervention*).tw,kf. |
|  |  | AND |
|  | Social Norms/ or Peer Influence/ or Peer Group/ | (incentiv* or default* or salien* or norm* or affect or messenger* or endors* or priming or remind* or commit* or ego* or feedback* or peer* or alert or fram* or play* or game or gamif*).tw,kf. |
|  |  | AND |
| String 2 | Exp Sanitation/ or exp Sanitary Engineering/ or Hand Disinfection/ or Soaps/ or exp Cross Infection/ or Hand Sanitizers/ or Social Distance/ or hygiene/ or hand hygiene/ | ((sanitation) or (hand adj1 (disinfection* or wash* or soap* or sanit* or hygiene*)) or (social distance*) or (avoid* adj3 (crowd* or contact)) or ((cover or etiquet*) adj2 (cough* or sneez*)) or (face mask*) or hygiene* or WASH or soap* or detergent* or water or toilet* or latrine* or borehole* or rainwater or (open adj1 (defecation or urination)) or sanitary).tw,kf. |
|  | Disinfection/ | (disinfect* or desinfect*).tw,kf. |
|  | exp Insect Control/ or Insect Repellents/ | (((Vector* or mosquito* or insect* or tick* or weed* or environmental or mechanical or biological or chemical) adj1 (control or management)) or IVM or repellent* or insecticide* or adulticide* or (insect growth regulator*) or predator* or trap* or (bed net*) or ((larv* or guppy) adj1 fish) or (cloth* adj2 (wear* or protect*)) or ((remov* or drain* or clea* or manage* or dispos* or eliminat*) adj2 (water* or trash* or waste or breed* or source*)) or (cover* adj2 (container* or food* or trash*))).tw,kf. |
|  | Exp pest control/ | ((animal* or rodent* or pest*) adj3 (control* or handl* or pet* or wild*)).tw,kf. |
|  | exp Food Handling/ or exp Food Safety/ | ((food* adj2 (hygiene or handl* or packing or storage* or prep* or cook* or dispens* or contaminat* or inspect* or safe* or Microbiology or Parasitology)) or cooking or (water or milk) adj2 (treatment or boil*) or "Hazard Analysis and Critical Control Points" or HACCP).tw,kf. |
|  | Tropical medicine/ or neglected diseases/ | NTD or NTDs or (neglected tropical disease*) or (infectious disease* of poverty) |
|  | Leishmaniasis/ | (leishman* or kala-azar or sandflies or Lutzomyia or phlebotomus).tw,kf. |
|  | Ascariasis/ or strongyloidiasis/ or  Ancylostomatoidea/ or  Trichuris/ | (ascari* or strongyl* or hookworm* or Ancylostoma or Ankylostoma or Necator or trichur*or whipworm* or soil-transmitted helminth* or geohelminth*).tw,kf. |
|  | Schistosomiasis/ | (schistosome* or schistomiasis or bilharz*).tw,kf. |
|  | Leprosy/ | (Lepros* or lepra or leprology or elephantiasis graecorum or hansen disease or hanseniasis or morbus Hansen or Mycobacterium leprae infection).tw,kf. |
|  | Elephantiasis, filarial/ | ((Filaria* adj2 (elephantiasis or lymphatic or lymphedema or lymphoedema or lymphooedema)) or Wuchereria bancrofti or wucherer* or brugia*).tw,kf. |
|  | Onchocerciasis/ | (onchocer* or river blindness).tw,kf. |
|  | Trachoma/ | (((Chlamydia* or granular) adj trachoma*) or trichiasis or trachoma*).tw,kf. |
|  | Rabies/ or rabies virus/ | (rabies or rabbia or (hydrophobia adj2 (agent or virus)) or lyssavirus* or Hubert disease).tw,kf. |
|  | Cysticercosis/ | (cysticercosis or neurocysticercosis or Taenia solium).tw,kf. |
|  | Dengue/ | (Dengue or dengue adj2 fever or shock syndrome or zika or chikungunya or *Aedes* or tiger mosquito*).tw,kf. |
|  | Chagas disease/ | (Chaga* adj2 disease*).tw,kf. |
|  | Echinococcosis/ | (echinococc* or (hydatid adj1 (diesease* or cyst*)) or multilocularis or granulosus or hydatid*).tw,kf. |
|  | Fascioliasis/ | (foodborne trematode* or clonorch* or opisthorch* or paragonim* or fasciolop* or fasciol* or intestinal fluke* or (Distomiasis adj1 hepatic) or Fasciol* or (liver fluke*) adj2 (disease* or infection*)).tw,kf. |
|  | trypanosomiasis, African/ | ((African or Gambian or Rhodesian or congo or negro) adj2 (trypanosom* or lethargy)) or sleeping sickness or (Trypanosoma brucei adj2 infection).tw,kf. |
|  | dracunculus nematodes/ | (Guinea worm* or dracontiasis or dracunculosis or Dracunculus).tw,kf. |
|  | Buruli ulcer/ | ((Buruli adj1 (ulcer* or disease*)) or Bairnsdale ulcer or Mycobacterium ulcerans).tw,kf. |
|  | Mycetoma/ or chromoblastomycosis/ | (chromoblastomycosis or chromomycosis or mycetoma or deep mycosis or Fonsecaea pedrosoi infection or Phialophora verrucosa infection).tw,kf. |
|  | Exp Ectoparasitic infestations/ | ((human ecto parasite*) or (*scaroptes scabei var hominis*) or (human scabies) or tungiasis or tunga penetrans or tunga trimamillata or (ectoparasite* adj2 (infection* or infestation* or disease*))).tw,kf. |
|  | Snake bites/ | (snake bite* or snake bite envenoming or snakebite*).tw.kf |

*Note.* MeSh: Medical SubjectsHeading; tw: tekst word; kf: keyword heading word
